# Cardioprotective effects of antiretroviral treatment in adolescents with perinatal HIV infection are heterogeneous depending on age at treatment initiation

**DOI:** 10.1101/2024.03.08.24303983

**Authors:** Itai M Magodoro, Carlos E Guerrero-Chalela, Brian Claggett, Stephen Jermy, Petronella Samuels, Landon Myer, Heather Zar, Jennifer Jao, Mpiko Ntsekhe, Mark J Siedner, Ntobeko AB Ntusi

## Abstract

The cardioprotective effects of antiretroviral treatment (ART) in adolescents with perinatal HIV infection (APHIV) may depend on age at ART initiation. We used cardiovascular magnetic resonance (CMR) to characterize and compare residual cardiac changes in apparently healthy APHIV with early and delayed ART initiation compared to sex- and age-similar HIV uninfected peers. We defined early and delayed ART as, respectively, treatment initiated at <5 years and ≥5 years of age. Cardiac function, mechanical deformation, geometry and tissue composition were assessed. APHIV had distinct albeit subclinical cardiac phenotypes depending on timing of ART initiation. For example, changes in early ART suggested comparatively worse diastology with preserved systolic function while delayed ART was associated with comparatively increased diffuse fibrosis and LV dilatation with reduced systolic function. The long-term clinical significance of these changes remains to be determined.

## Introduction

Cardiovascular morbidity and mortality were common among adolescents with perinatal HIV infection (APHIV) prior to wide availability of antiretroviral therapy (ART) [1], [2]. Successful treatment of HIV infection has been associated with significant improvements in cardiovascular status of these young people, especially those in high-income countries (HICs), confirming the cardioprotective effects of ART [3]. In contrast, subclinical cardiac abnormalities remain prevalent in their peers in sub-Saharan Africa (SSA) [4], [5], [6] where 90% of global pediatric HIV patients reside [7]. Here, pediatric ART coverage is neither universal (54% in 2020) [8]^9^ nor is it started early (median age at initiation 7.9 years (2018)) [7], [8] before substantial immunosuppression and cardiac damage set in [3]. The concern is that these cardiac abnormalities in early life will likely be a substrate for symptomatic cardiovascular dysfunction in adulthood, particularly as maturing APHIV are cumulatively exposed to traditional cardiovascular risk factors [9]. Compounding this is the reality that perinatal HIV transmission in SSA is not expected to be eliminated before 2030 [10] meaning that even modestly increased relative risks of cardiovascular disease will multiply into a large absolute case burden as more APHIV survive into adulthood.

However, our knowledge of the phenotypes and epidemiology of these perinatal HIV infection (PHIV) related cardiac forms remains incomplete, especially regarding the timing of ART. This, in turn, curtails our capacity to intervene preventively, let alone selectively. One major drawback to cardiovascular research in APHIV in SSA has been its almost exclusive reliance on echocardiography [4], [5], [6] notwithstanding the modality’s limitations [11]. Cardiovascular magnetic resonance (CMR), with its multi-parametric capabilities, is the gold standard for cardiac volumetric and functional assessment [11]. Because of its capabilities to characterize myocardial tissue, CMR has been likened to a “virtual biopsy” [12] and is thus well suited to the challenge at hand. Unfortunately, CMR is inaccessible in the typical adolescent HIV care setting in SSA [13]. In response to these gaps, we conducted a CMR study to comprehensively characterize residual cardiac abnormalities in ART-experienced APHIV in Cape Town, South Africa. Our specific objective was to identify and quantify domain specific, i.e., geometry, systolic and diastolic function, mechanical deformation, and tissue composition, cardio-protective effects of early versus delayed ART in a high PHIV burden African setting.

## Methods

We followed the guidelines of the Strengthening the Reporting of Observational Studies in Epidemiology (STROBE) in the conduct and reporting of our analyses [14].

### Study population and setting

This was an observational cross-sectional study of participants enrolled in the ongoing prospective Cape Town Adolescent Antiretroviral Cohort (CTAAC). The CTAAC, described fully elsewhere [5], enrolled young persons, then aged 9-14 years old (2012-2013), with PHIV and established on ART and a comparison group of similar age and sex HIV uninfected peers. CTAAC’s overall aim is to track the development of chronic diseases in children and adolescents with PHIV. For the current analysis, we approached and enrolled cohort members consecutively presenting for a scheduled study visit (Visit #9, 2018-2019) to complete a comprehensive CMR examination. Participants were eligible to participate if they were aged ≥13 years, had no history of clinical heart disease, no active systemic infections, no current use of anti-inflammatory therapies, and no known contraindications to CMR [e.g., gadolinium sensitivity or reduced glomerular filtration rate (≤30ml/min/1.73m^2^)].

### Ethics

The investigation conforms with the principles outlined in the Declaration of Helsinki. The Human Research Ethics Committee of the Faculty of Health Sciences, University of Cape Town, approved all study activities. Participants’ and parents’ informed assent and consent were obtained in accordance with South African practice.

### Data collection

Sociodemographic data, past medical history, age at ART initiation, WHO HIV clinical staging (I-IV) and CD4+ cell count at time of HIV diagnosis (presumed *nadir CD4+*) and ART exposure up to the time of CMR imaging were collected under the parent CTAAC protocol. Seated brachial blood pressure (BP) and anthropometrics were measured. The last two of three same arm BP readings were averaged to give final systolic (SBP), diastolic (DBP), and mean arterial (MAP) blood pressure. Height, weight, and body mass index (BMI) were referenced to age and sex according to WHO growth reference data,[15] to obtain height-for-age (HFA), weight-for-age (WFA), and BMI-for-age (BFA) z scores. Stunting (HFA z score<-2), underweight (WFA z score <-2), wasting (BFA z score <-2) and overweight/obesity (BFA z score >1) were also determined.

Venous blood was collected from APHIV for measurement of current CD4+ cell count (Beckman Coulter FC500 MPL analyzers, USA) and HIV viral load (lower limit of detection 40 copies/mL; Roche Cobas AmpliPrep/TaqMan). We classified HIV-related markers into the following ordinal categories: age at ART initiation (early: <5 years old, and delayed: ≥5 years old); nadir and current CD4+ (<350 vs. ≥350 cells/mL); HIV viral load (viremia: ≥40 vs. aviremia: <40 copies/mL); AIDS history (HIV clinical stage IV vs. stages I-III); and ART history (protease inhibitor (PI) exposure vs. none; and non-nucleoside reverse transcriptase inhibitor (NNRTI) exposure vs. none). ART exposure duration was measured from initiation to time of CMR examination.

### Cardiovascular magnetic resonance (CMR) procedures Image acquisition

As previously described, protocol-directed CMR was performed at 3 Tesla (Siemens, Magneton Skyra, Erlangen, Germany) [16]. The protocol included balanced steady-state free precession (bSSFP) for volumetrics and function, T1 mapping by Modified Look-Locker Inversion Recovery (MoLLI) sequence, T2 mapping using fast-low-angle-shot (FLASH) sequence, and late gadolinium enhancement (LGE) using a phase sensitive inversion recovery (PSIR) sequence 10-15 minutes after gadolinium contrast administration. Imaging parameters for the sequences used were maintained between participants and are summarized in **Supplementary Table S1**. CMR image analysis was done offline, blind to PHIV/ART status, using proprietary software (CVI42, Circle Cardiovascular Imaging, Calgary, Canada) [17].

### Image analysis

LV end-diastolic volume (LVEDV), LV end-systolic volume (LVESV), LV ejection fraction (LVEF) and LV mass (LVM) were automatically derived from SAX views. Relative wall thickness (RWT) was calculated as twice inferolateral wall thickness divided by LV end-diastolic diameter, whereby both inferolateral wall thickness and LV end-diastolic dimension were measured in end-diastole from a SAX frame immediately basal to the tip of papillary muscle tips [17]. We assessed the tricuspid annular plane systolic excursion (TAPSE) in the LAX four-chamber view by measuring the distance travelled by the tricuspid annulus from end-diastole to end-systole. Basal, mid-ventricle, and apical SAX slices were manually contoured to outline the endocardium and epicardium, and generate maps for native T1, T2 and extracellular volume (ECV) estimation. We calculated the T2 signal intensity (SI) ratio as previously described [17], and visually assessed the presence or absence of LGE. LV circumferential, radial, and longitudinal deformation parameters (global systolic strain, time to peak systolic strain, peak diastolic strain rate, and diastolic velocity) were analyzed by semi-automated feature tracking during diastole and systole from long axis (LAX) and SAX cine images.

### Statistical analyses

A formal sample size calculation was not performed due to the exploratory nature of the study. Missingness in the key study variables was less than 2%, and thus we proceeded with complete case analysis. Where relevant, cardiovascular parameters were indexed to Mosteller body surface area (BSA) [18] to account for somatic growth, and are indicated by the postscript (i). Ninety-five percent confidence intervals (95%CI) were calculated based on bootstrap replications. Summary statistics are presented as mean (standard deviation, SD) or median (interquartile range, IQR) for continuous variables and number (percent) for categorical variables. Our primary exposure groups were HIV uninfected, APHIV with early ART, and APHIV with delayed ART. Comparisons by exposure were made using linear and quintile regression modeling, as appropriate. Differences in the distribution of blood pressure and standardized anthropometric measures were visually displayed.

There are currently no published reference standards for CMR-derived parameters for children and adolescents (<20 years old) in SSA. We, therefore, derived z scores of cardiac variables for APHIV by referencing their HIV uninfected peers as normative. For a given variable, its z score was calculated by: (mean*_aphiv_* – mean*_hiv uninfected_*)/(SD*_hiv uninfected_*). A z-score measures in SD units the distance of a raw score from the mean. Thus, a z score of 0.5 (or -0.5), for example, means that APHIV have a cardiac parameter that is 0.5 SDs above (or below) the HIV uninfected (i.e., control) group’s mean. To quantify group differences in cardiac parameters, we classified the z scores as small (<0.5), medium (≥0.5 and <0.8), or large (≥0.8) using Cohen’s criteria [19]. We fitted linear regression models to evaluate the association between PHIV/ART status and cardiac parameters (measured as z scores) after adjustment for age, sex, MAP and BMI. All statistical analyses were performed using Stata version 17.0 (StataCorp, College Station, TX) and R, version 3.6.3 (R Foundation for Statistical Computing, Vienna, Austria).

## Results

### Characteristics of study sample

We enrolled 103 [mean (SD) age 15.1 (1.7) years; male sex 49 (48%)] adolescents to undergo CMR and blood collection from the 422 CTAAC participants [335 (79%) PHIV] in active follow-up at visit 9. Of these, 37 were HIV uninfected, 45 were APHIV with early ART initiation and 21 APHIV with delayed ART initiation (**Table 1**). Both absolute (**Table 1**) and age-standardized (**Figure 1A-C**), weight, height and BMI were largely comparable across the three groups. However, stunting was more common among APHIV with delayed (24%) and early ART (18%) than among HIV uninfected (8%) counterparts. Overweight/obesity was, on the other hand, more frequently observed among HIV uninfected participants (38%) than among APHIV with either early (16%) or delayed (11%) ART initiation (**Table 1**). Other than systolic BP, which was higher among HIV uninfected participants [median (IQR): 108 (103, 113) mmHg] than APHIV with delayed ART [101 (98, 110) mmHg; p = 0.040], there were no significant group differences either diastolic BP or MAP (**Figure 1D-F**).

**Figure 1.**
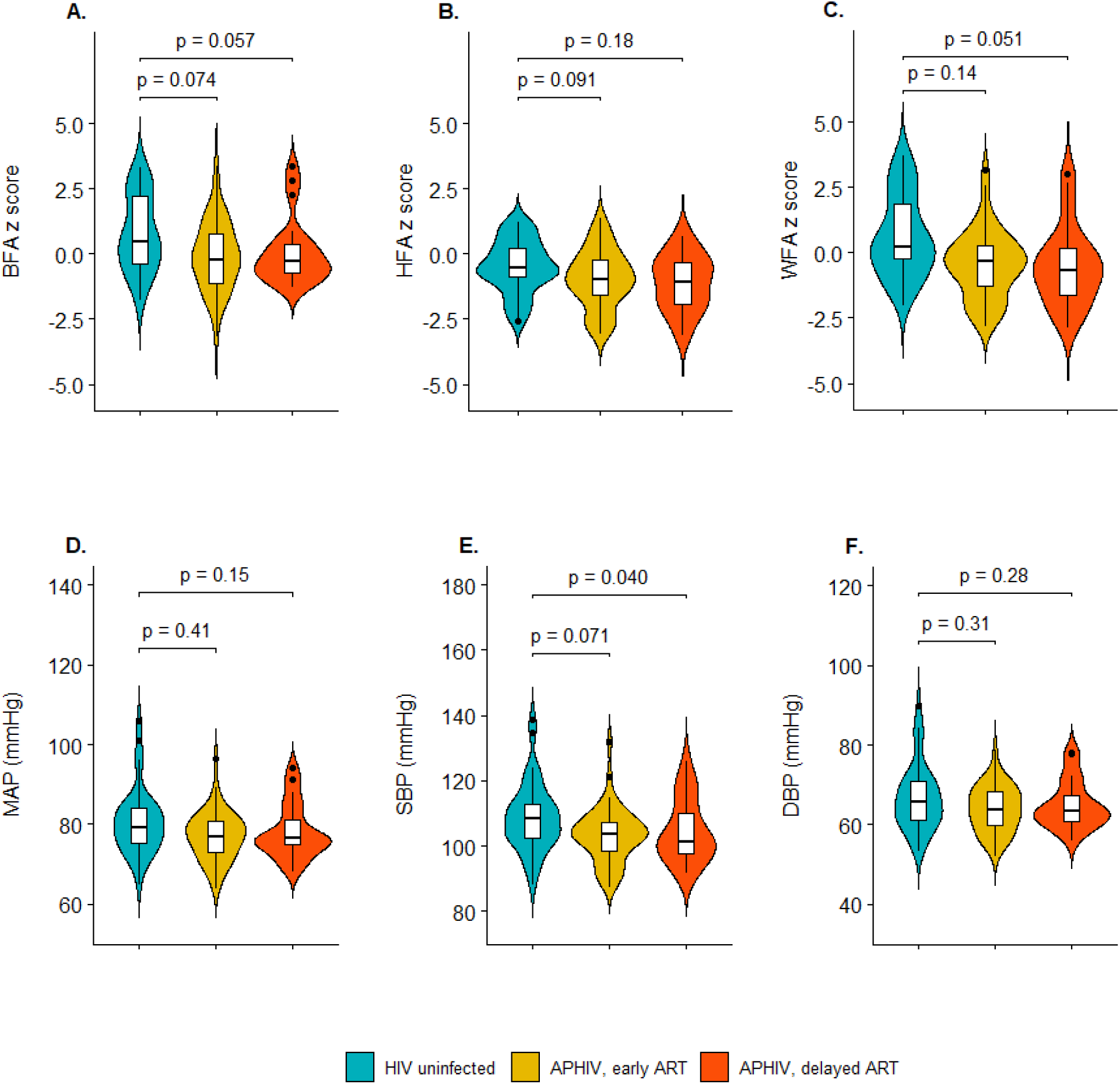
Distribution of age-standardized anthropometric measures and blood pressure according to perinatal HIV infection and antiretroviral treatment status. P-values are estimated from unadjusted median quintile regression models referenced to HIV uninfected group. BFA = BMI-for-age; HFA = height-for-age; WFA = weight-for-age; MAP = mean arterial blood pressure; SBP = systolic blood pressure; DBP = diastolic blood pressure.

**Table 1.** Participants’ baseline characteristics according to perinatal HIV infection and antiretroviral treatment status.

| Characteristic | PHIV and ART Status |  |  |  |
| --- | --- | --- | --- | --- |
|  | HIV uninfected | APHIV |  |  |
|  |  | Early ART initiation | Delayed ART initiation | Total |
| Number | 37 | 45 | 21 | 66 |
| *Sociodemographic* |  |  |  |  |
| Mean age (years) | 15.0 (1.8) | 15.0 (14.0, 16.0) | 15.3 (1.6) | 15.3 (1.6) |
| Male sex | 15 (41%) | 23 (51%) | 11 (52%) | 34 (52%) |
| *Anthropometric*<sup>a</sup> |  |  |  |  |
| BSA(m<sup>2</sup>) | 1.6 (0.2) | 1.5 (0.2) | 1.5 (0.2) | 1.5 (0.2) |
| Height (cm) | 159 (8) | 157 (9) | 159 (10) | 158 (9) |
| Weight (kg) | 60.7 (17.2) | 50.8 (12.9) | 55.1 (15.0) | 52.2 (13.6) |
| BMI (kg/m<sup>2</sup>) | 23.9 (6.6) | 20.6 (4.6) | 21.8 (5.7) | 20.9 (5.0) |
| Stunted | 3 (8%) | 8 (18%) | 5 (24%) | 13 (20%) |
| Underweight | 1 (3%) | 5 (11%) | 3 (14%) | 8 (12%) |
| Wasted | 0 (0%) | 3 (7%) | 0 (0%) | 3 (5%) |
| Overweight/Obese | 14 (38%) | 7 (16%) | 2 (11%) | 9 (15%) |

Values are reported as mean (SD)or number (%).
<sup>a</sup> Stunted = height-for-age z score < -2; Underweight = weight-for-age z score < -2; Wasted = BMI-for-age z score < -2; Overweight/obese = BMI-for-age z score >1.

### HIV-related characteristics

Median (IQR) age at ART initiation was 2.4 (0.9, 3.4) years for APHIV with early ART versus 7.2 (6.7, 8.2) years (p<0.001) for those with delayed ART (**Table 2**). Although treatment with PIs (yes/no) was equally frequent in early (51%) and delayed ART (48%; p=0.74) APHIV at time of CMR, those with early ART had considerably longer cumulative PI exposure [5.1 (0.0, 10.3) years] than those with delayed ART [0.1 (0.0, 4.9) years; p<0.001]. There were no significant group differences with respect to current ART regimen, which included a backbone of two nucleoside reverse transcriptase inhibitors (NRTIs) plus either one PI or one NNRTI agent. However, compared to early ART initiation, delayed ART was associated with a greater degree of immunosuppression [nadir CD4+ cell count: 457 (368, 720) vs. 547 (422, 835) cells/mL; p=0.025], and a lower degree of immune reconstitution [current CD4+ count: 592 (402, 795) vs. 753 (558, 928) cells/mL; p=0.011].

**Table 2.** HIV disease markers stratified by antiretroviral treatment status among participants with perinatal HIV infection.

| Characteristic | APHIV with |  | P value |
| --- | --- | --- | --- |
|  | Early ART initiation | Delayed ART initiation |  |
| Number | 45 | 21 |  |
| Age at ART initiation (years) | 2.4 (0.9, 3.4) | 7.2 (6.7, 8.2) | <0.001 |
| Overall ART duration (years) | 12.8 (11.3, 13.8) | 8.1 (7.8, 9.7) | <0.001 |
| Cumulative NRTI exposure (years) | 12.8 (11.3, 13.8) | 8.1 (7.8, 9.7) | <0.001 |
| Cumulative PI exposure (years) | 5.1 (0.0, 10.3) | 0.1 (0.0, 4.9) | 0.015 |
| Cumulative NNRTI exposure (years) | 7.0 (3.4, 10.6) | 6.4 (3.7, 8.1) | 0.49 |
| Cumulative INSTRI exposure (years)<sup>α</sup> | 0 | 0 | 0.92 |
| Current ART regimen |  |  |  |
| 2 different NRTIs + 1 NNRTI | 21 (47%) | 11 (52%) | 0.74 |
| 2 different NRTIs + 1 PI | 23 (51%) | 10 (48%) |  |
| Other<sup>β</sup> | 1 (2%) | 0 (0%) |  |
| Undetectable HIV RNA viral load<sup>δ</sup> | 28 (64%) | 12 (60%) | 0.78 |
| Current CD4+ cell count (cells/mL) | 753 (558, 928) | 592 (402, 795) | 0.011 |
| <350 | 5 (12%) | 3 (14%) | 0.76 |
| Nadir CD4+ cell count, (cells/mL) | 574 (422, 835) | 457 (368, 720) | 0.025 |
| <350 | 7 (16%) | 5 (24%) | 0.44 |
| History of AIDS-related illness | 12 (29%) | 4 (21%) | 0.54 |

Values are reported as median (25<sup>th</sup>, 75<sup>th</sup> percentile) or number (%).
<sup>α</sup> Only one participant exposed to an integrase strand transfer inhibitor for total duration of 1.7 years.
<sup>β</sup> Participant receiving 1 NRTI + 1 PI + 1 INSTI.
<sup>δ</sup> Detection limit ≥40 viral copies/mL.
PI = protease inhibitors; NNRTI = nonnucleoside reverse transcriptase inhibitors; NRTI = nucleoside reverse transcriptase inhibitors; INSTI = integrase strand transfer inhibitor.

### Left ventricular structure and function

**Table 3** summarizes absolute mean values of left ventricular (LV) indices while **Figures 2** and **3** report the corresponding z score measures. Compared to HIV uninfected participants, APHIV with early ART initiation had reduced LVMi [adjusted mean (95%CI) z score: -0.36 (-0.61, -0.11); p = 0.042] but similar LV volumes, e.g., LVEDVi [adjusted z score: 0.19 (-0.13, 0.51); p = 0.25] and RWT [adjusted z score:

**Figure 2.**
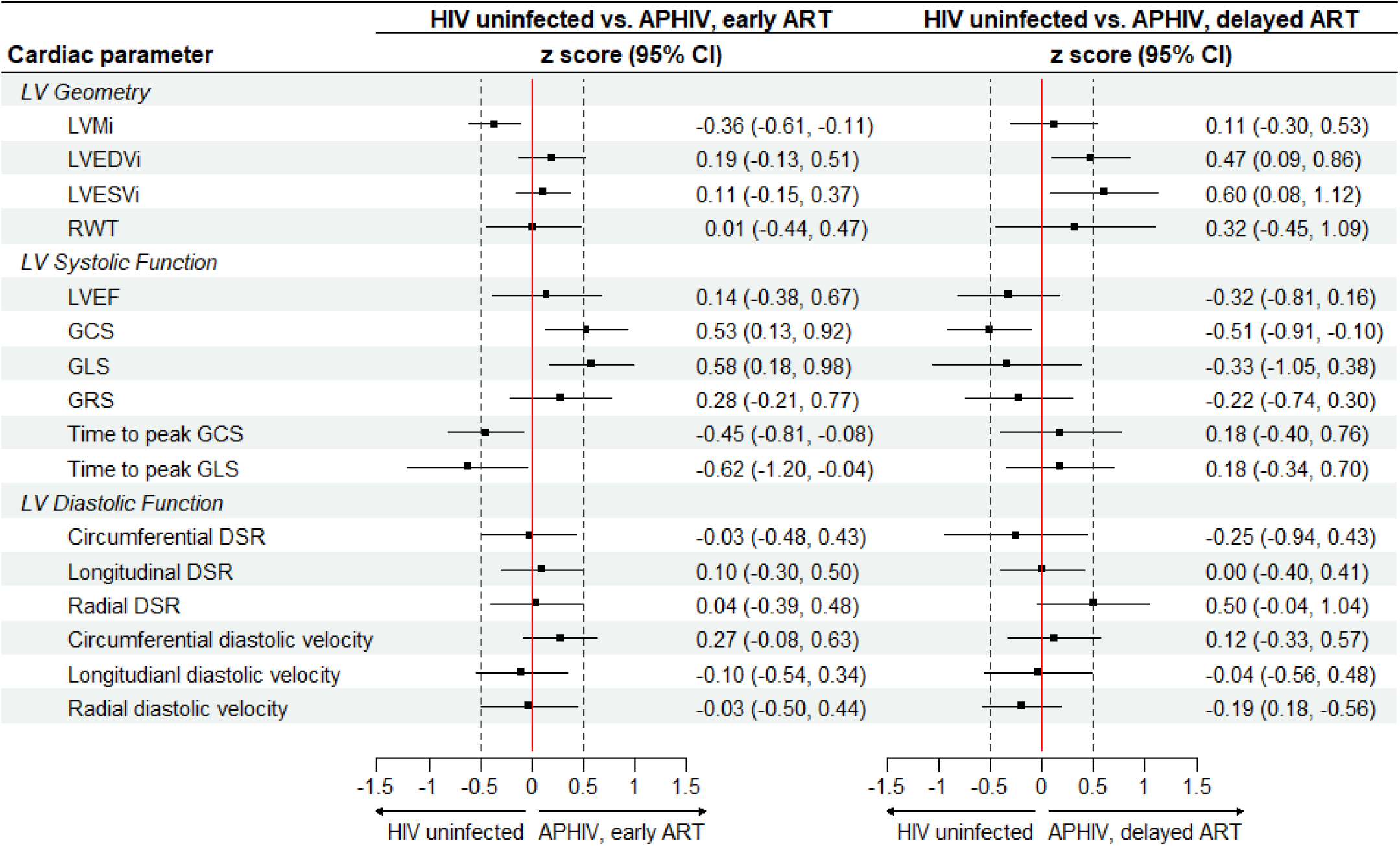
z scores of left ventricular parameters according to antiretroviral treatment status among participants with perinatal HIV infection. Abbreviations as elsewhere defined. Postscript (i) = indexed to body surface area.

**Figure 3.**
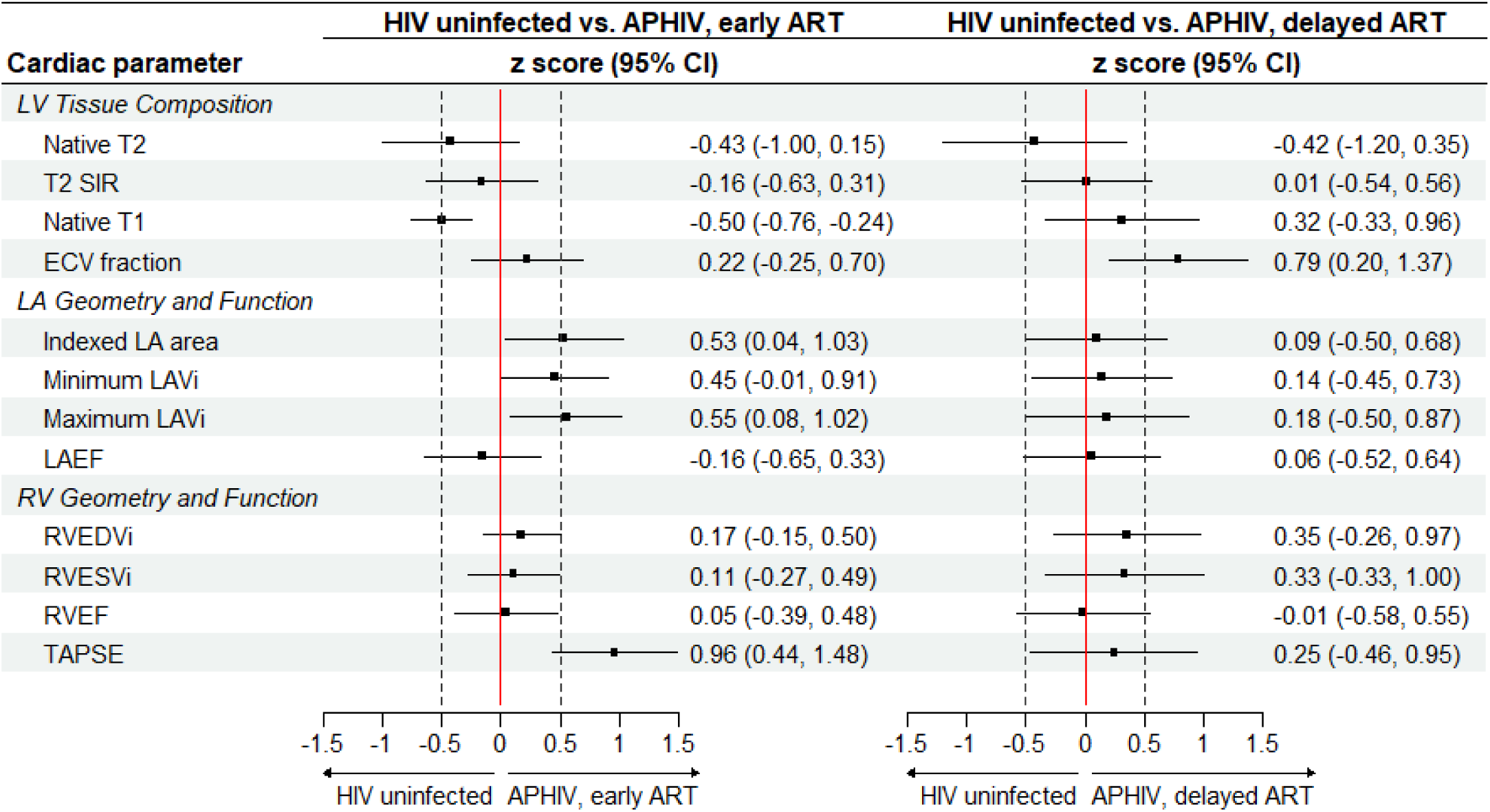
z scores of left and right ventricular and left atrial parameters according antiretroviral treatment status among participants with perinatal HIV infection. Abbreviations as elsewhere defined. Postscript (i) = indexed to body surface area.

0.01 (-0.44, 0.47); p = 0.96]. They also had comparatively better LV systolic function when assessed by mechanical deformation indices like GCS [z score: 0.53 (0.0.13, 0.92); p = 0.023] and time to peak GCS [adjusted z score: -0.45 (-0.81, -0.08); p = 0.041]. However, the two groups had similar LV diastolic function measured by either diastolic strain rate or diastolic velocity. (**Figure 2**).

On the other hand, preserved LVMi [adjusted z score: 0.11 (-0.30, 0.53); p = 0.59] and RWT [adjusted z score: 0.32 (-0.45, 1.09); p = 0.34] in APHIV with delayed ART were accompanied by a relatively dilated LV chamber [LVEDVi adjusted z score: 0.47 (0.0.09, 0.86); p = 0.014]. Although individually not achieving statistical significance (except GCS [adjusted z score: -0.51 (-0.91, -0.10); p = 0.020]), all study measures of LV systolic function (ejection fraction, systolic strain and time to peak systolic strain) were collectively suggestive of reduced function in APHIV with delayed ART compared to their HIV uninfected peers. However, LV diastolic function was preserved as determined by measures like longitudinal [adjusted z score: -0.04 (-0.45, 0.73); p = 0.98] and radial [adjusted z score: 0.20 (-0.34, 0.57); p = 0.61] diastolic strain rate (**Figure 2**). The distribution of absolute means (**Table 3**) was largely consistent with the adjusted z score based results. Of note, all the observed PHIV/ART related LV differences were of small-to-moderate size based on the z scores.

### Left atrial structure and function

Maximum LAVi was 13.8 (12.0, 15.6) mL/m^2^ for HIV uninfected controls compared to 14.8 (12.3, 17.3) mL/m^2^ (p = 0.56) for APHIV with early ART and 16.0 (14.5, 17.5) mL/m^2^ (p = 0.056) for APHIV with delayed ART (**Table 3**).

After adjustment, early ART [maximum LAVi adjusted z score: 0.55 (0.08, 1.02); p = 0.032], and not delayed ART [maximum LAVI adjusted z score: 0.28 (-0.40, 0.97); p = 0.36], was associated with larger LA chambers (**Figure 3**). The magnitude of differences in LA chamber dimension associated with early ART were of moderate size. There were no significant differences in LAEF with or without adjustment for either early ART [adjusted z score: -0.16 (-0.65, 0.33); p = 0.47] or delayed ART [LAEF adjusted z score: 0.06 (-0.52, 0.64; p = 0.82].

### Right ventricular structure and function

The RVEDVi was 77.0 (71.4, 82.5) mL/m^2^ for HIV uninfected controls compared to 83.0 (78.9, 87.0) mL/m2 (p = 0.099) for APHIV with early ART and 87.0 (78.1, 95.8) mL/m^2^ (p = 0.029) for APHIV with delayed ART (**Table 3**). These corresponded to adjusted z scores of 0.27 (-0.05, 0.60) for APHIV with early ART and 0.35 (-0.26, 0.97) for APHIV with delayed ART. Thus, while there were no statistically significant PHIV/ART-related differences in RV chamber size or RVEF, APHIV with early ART had considerably improved RV systolic function if assessed by TAPSE [adjusted z score: 0.96 (0.44, 1.48); p<0.001].

### Tissue characteristics

There were no significant differences in T2 mapping and T2-STIR signal intensity ratios between HIV uninfected participants and APHIV with early or delayed ART, whether measured in absolute values (**Table 3**) or as adjusted z scores (**Figure 3**). However, APHIV with delayed ART had a greater ECV fraction [29.5 (28.0, 31.0) %; p = 0.044] than, while APHIV with early ART had a comparable ECV fraction [28.7 (27.7, 29.7) %; p = 0.13] to HIV uninfected controls [28.1 (27.0, 29.1) %]. This translated to adjusted z scores of 0.79 (0.20, 1.37) (p = 0.010) for delayed ART and 0.32 (-0.15, 0.80) (p = 0.17) for early ART (**Figure 3**).

## Discussion

In this CMR study, we identified distinct albeit subclinical cardiac phenotypes associated with early (< 5 years of age) and delayed (≥5 years of age) ART initiation among APHIV. Changes seen with early ART included relatively low LV mass, improved LV systolic function, and LA enlargement notwithstanding preserved LV diastolic mechanical deformation. On the other hand, delayed ART was associated with comparative LV chamber enlargement, reduced systolic function and greater interstitial fibrosis. These differences may be rooted in differing degrees of systemic inflammatory activation and immunosuppression correlated with ART timing in PHIV and, in turn, the extent and type of cardiac damage sustained in early life. Whether and how these difference might impact clinical cardiovascular trajectories of APHIV initiating ART at different ages remains to be determined.

Our results, while consistent with previous reports of cardioprotection with ART in APHIV, also highlight the potential heterogeneity of these effects. This may have far-reaching clinical implications in the setting of SSÀs pediatric HIV epidemic. Currently available studies [3], [5], [6], [20] of ART and the heart in Africàs young persons with PHIV have almost exclusively included participants with late HIV diagnoses, delayed ART initiation, and/or short duration of ART exposure. These studies found not infrequent cardiac functional and morphological abnormalities including LV hypertrophy (67% prevalence [21]), dilatation (8% prevalence [1]), systolic (5.5% - 33% prevalence [6], [21]) and diastolic (36% prevalence [22]) dysfunction, and RV dilatation (29% [21]), and often accompanied by symptoms of exertional dyspnea, leg swelling and cough, among others. In contrast, Mahtab *et. al.,* (2020) [5] reported very low prevalence of echocardiographic abnormalities (6.7% LV hypertrophy, 7.6% diastolic dysfunction, 0.2% systolic dysfunction) in a South African APHIV cohort. This is possibly the only African report to date of a cohort with early ART initiation and prolonged ART exposure. Their results are in keeping with studies in HICs where pediatric ART is both universal and early [3], [23], [24].

Our study did not enumerate cardiac abnormalities, and thus we cannot compare their prevalence with prior reports. However, there are notable differences in cardiac phenotype observed with either early or delayed ART in our study compared to previous reports. For example, delayed ART in our cohort was phenotypically marked by relative LV chamber enlargement, reduced systolic function and greater interstitial fibrosis. LV diastolic function was preserved as were LA and RV function and morphology. In other studies, LV enlargement and reduced LV systolic function with delayed ART were reported along with other abnormalities including LV hypertrophy, impaired LV diastolic function, RV dilatation and increased pulmonary artery systolic pressure [1], [4], [6], [20], [21]. Further, the cardiac changes seen with delayed ART initiation in our study were not associated with any symptoms and were, at best, of moderate size. This contrasts with other reports, mostly from SSA, where cardiac abnormalities in delayed ART frequently occurred with symptoms of heart failure. It is noteworthy that ART duration (median 8.1 years) with delayed ART in our cohort was much longer than in comparable studies.

Similarly, studies from HICs invariably found no clinically significant cardiac differences between those with PHIV and early ART initiation and their HIV uninfected counterparts [3], [23], [24]. The subclinical differences that have been described, at times contradictory, entail LV systolic strain, LV fractional shortening, and LV mass. Our observation of reduced LV mass with early ART echoes the results of a US study of children and adolescents early exposed to ART [25]. Previously unreported, however, was our finding of relative LA enlargement. LA enlargement is often a marker of chronically elevated LA pressure, and signifies increased LV stiffness and diastolic dysfunction. However, formal LV diastolic functional assessment requires measurement of flow and velocities at the mitral annulus, mitral inflow and pulmonary veins [26] - which we did not evaluate - together with LA dimensions.

Nonetheless, the cardiac phenotype we observed in APHIV with early ART, *i.e*., preserved systolic function with likely impaired diastology, may mirror the shifts in epidemiology of HIV cardiomyopathy that have been observed in adult HIV care in HICs [27], [28]. LV diastolic dysfunction, often with preserved systolic function, is now described in one in two (43-50%) [27], [28] asymptomatic HIV infected adults on long-term suppressive ART. This in turn warrants concern about future heart failure (HF) risks, especially HF with preserved ejection (HFpEF). HFpEF is often preceded by LV diastolic dysfunction with normal systolic function and is the predominant manifestation of HF in adults with HIV. It is noteworthy that uptake of early infant diagnosis of HIV and linkage to pediatric ART are increasing across SSA [29]. Thus, the prevalence and incidence of the cardiac phenotype associated with APHIV with early ART are likely to increase. Follow-up studies mapping the evolution and long-term outcomes of this cardiac phenotype are, therefore, urgently required.

### Strengths and limitations

We believe that our study is among the first to deploy CMR in adolescents with PHIV in SSA. Previous studies have invariably used echocardiography, which despite its lower cost, ready availability, and ease of use, has well documented limitations in accuracy and reproducibility. CMR, in contrast, has superior spatial resolution, excellent inter- and intra-study reproducibility, no acoustic window dependency, and makes no geometric assumptions. A drawback, however, was lack of repeat imaging precluding us from knowing whether cardiac status was static, improving, or deteriorating. Similarly, the study’s cross-sectional design prevents causal inferences. Our small sample size was another limitation as it prevented examination of important subgroups like sex, and the estimation of precise results. Lastly, our HIV uninfected participants may not have been the ideal reference group given their comparatively high rates of overweight/obesity, and the potential for confounding. However, this limitation will continue to plague adolescent cardiac imaging research in Africa until normative CMR ranges are published.

## Conclusions

We found residual albeit subclinical cardiac changes in South African APHIV on successful antiretroviral therapy suggesting that ART is cardioprotective. However, these cardioprotective effects are heterogenous and may depend on age ART initiation. Their long-term clinical significance remains to be determined, especially considering growing numbers of APHIV who are surviving into adulthood in Africa.

## Data Availability

All data produced in the present study are available upon reasonable request to the authors.

## Acknowledgements

In addition to thanking all study participants and their families, the authors would also like to acknowledge the support of the following people who made the study possible: Sana Mahtab, Nana Akua Asafu-Agyei, Landiwe Daka, Sharon Wakefield, Shanaaz Davids, Nikelwa Mvango-Njemla, Nomawethu Jele, Nomandla Udogwu*, Pumeza Nazo, Njemia Nikelwa, Keyola George, Akhona Mazingi, Tafadzwa Mautsa, Mothabisi Nyathi, Faith Qeja, Luyanda Nkondlwana, Mazwi Maishi, Mariaan Jaftha, Daniel Doetz, Liezl Julius, Michael Wheeler, Shaheed Sorathia, Nabeal Kaskar, Dave Nshuntishema, Okeyo, Patricie Niyitegeka, and Ben Jann.

*posthumously.

## Supplementary Material

**Supplementary Table S1.**
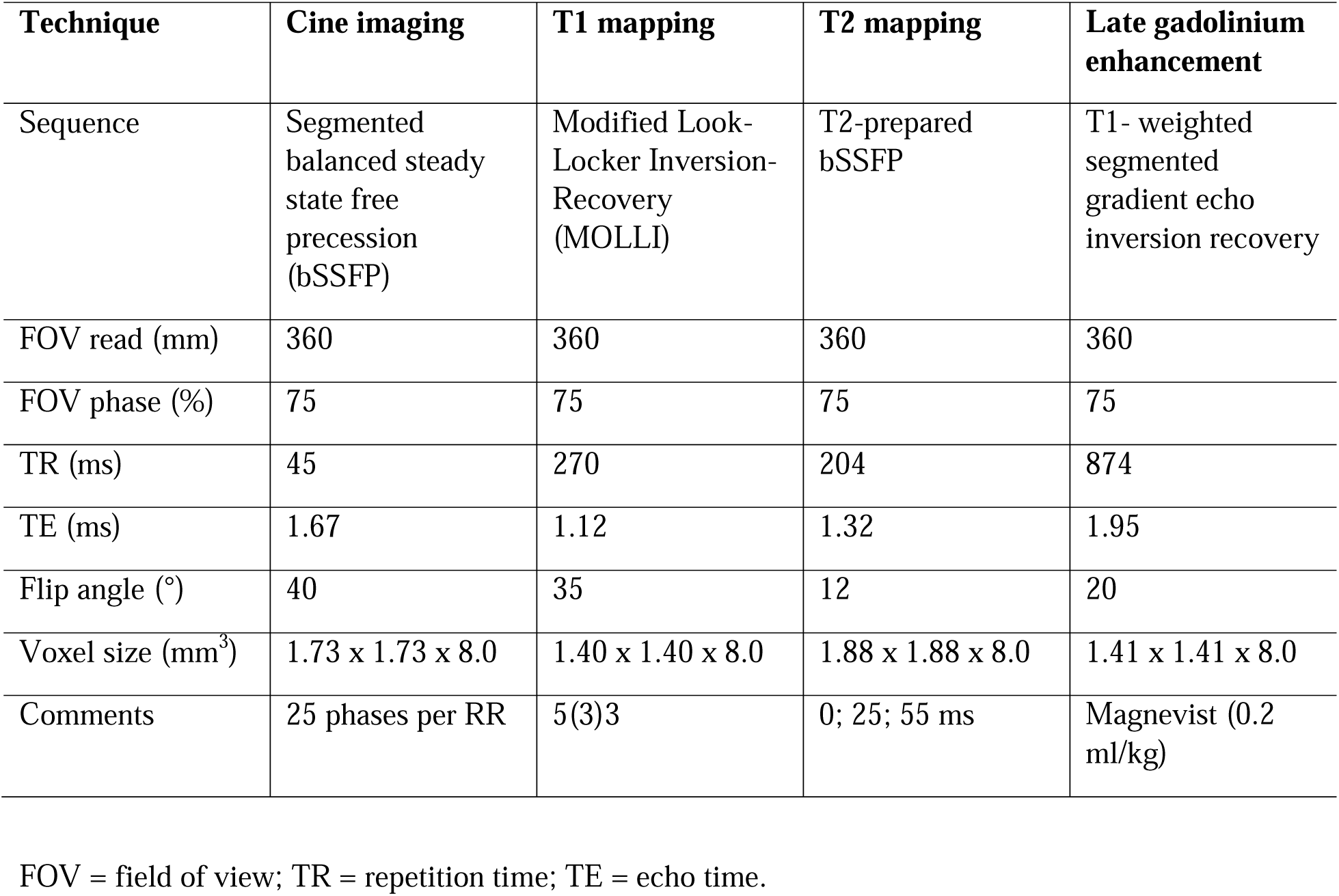
Cardiac magnetic resonance (CMR) sequence parameters.

