## Supplementary material for "Cardioprotective effects of antiretroviral treatment in adolescents with perinatal HIV infection are heterogeneous depending on age at treatment initiation": Table 3

**Table 3. Participants' absolute mean values of cardiac parameters stratified by perinatal HIV infection and antiretroviral treatment status.**

| **Cardiac parameter** | **PHIV and ART Status** | | | | |
| --- | --- | --- | --- | --- | --- |
|  | **HIV uninfected** | **APHIV with early ART initiation** | | **APHIV with delayed ART initiation** | |
|  | **Mean (95% CI)** | **Mean (95% CI)** | **P value**^α^ | **Mean (95% CI)** | **P value**^β^ |
| Number | 37 | 45 |  | 21 |  |
| ***LV Geometry*** |  |  |  |  |  |
| LVMi (g/m^2^) | 58.5 (54.6, 62.5) | 54.9 (52.1, 57.8) | 0.22 | 59.1 (55.5, 62.7) | 0.83 |
| LVEDVi (mL/m^2^) | 77.5 (73.4, 83.5) | 81.3 (77.7, 85.0) | 0.12 | 86.7 (78.9, 94.5) | 0.028 |
| LVESVi (mL/m^2^) | 28.9 (26.6, 31.2) | 30.5 (28.1, 32.8) | 0.26 | 34.5 (30.7, 38.2) | 0.019 |
| Relative wall thickness | 0.34 (0.32, 0.36) | 0.34 (0.33, 0.35) | 0.95 | 0.36 (0.33, 0.38) | 0.19 |
| ***LV Systolic Function*** |  |  |  |  |  |
| LVEF (%) | 63.0 (61.5, 64.6) | 63.0 (60.9, 65.0) | 0.83 | 60.2 (58.5, 62.0) | 0.18 |
| Peak systolic strain (%) |  |  |  |  |  |
| Circumferential | -20.1 (-21.1, -19.1) | -21.5 (-22.4, -20.6) | 0.020 | -20.0 (-22.2, -19.8) | 0.23 |
| Longitudinal | -15.7 (-16.3, -15.1) | -16.9 (-17.6, -16.2) | 0.041 | -16.1 (-17.2, -14.9) | 0.72 |
| Radial | 33.2 (31.3, 35.1) | 34.3 (32.4, 36.3) | 0.35 | 32.5 (30.1, 34.9) | 0.52 |
| Time to peak systolic strain (ms) |  |  |  |  |  |
| Circumferential | 247.4 (185.0, 309.8) | 205.3 (158.9, 251.7) | 0.56 | 227.8 (142.5, 313.1) | 0.88 |
| Longitudinal | 347.6 (333.0, 362.2) | 316.3 (293.0, 339.7) | 0.034 | 343.0 (328.6, 357.3) | 0.91 |
| ***LV Diastolic Function*** |  |  |  |  |  |
| Peak diastolic strain rate (s^-1^) |  |  |  |  |  |
| Circumferential | 1.41 (1.34, 1.48) | 1.40 (1.34, 1.47) | 0.86 | 1.33 (1.23, 1.44) | 0.20 |
| Longitudinal | 1.09 (1.00, 1.18) | 1.12 (1.02, 1.22) | 0.67 | 1.05 (0.93, 1.17) | 0.63 |
| Radial | -2.60 (-2.79, -2.42) | -2.58 (-2.74, -2.43) | 0.85 | -2.28 (-2.47, -2.10) | 0.029 |
| Peak diastolic velocity (cm/s) |  |  |  |  |  |
| Circumferential | 11.7 (5.4, 17.9) | 16.1 (12.8, 19.4) | 0.17 | 14.8 (9.1, 20.4) | 0.44 |
| Longitudinal | 28.8 (25.9, 31.7) | 27.1 (24.7, 29.4) | 0.37 | 27.5 (23.1, 32.0) | 0.60 |
| Radial | 38.5 (36.5, 40.5) | 37.9 (36.1, 39.6) | 0.65 | 36.8 (33.5, 40.0) | 0.31 |
| ***LV Tissue Composition*** |  |  |  |  |  |
| Native T2 (ms) | 38.8 (38.3, 39.3) | 38.4 (37.7, 39.1) | 0.16 | 38.5 (37.4, 39.5) | 0.58 |
| T2 SIR | 1.59 (1.49, 1.69) | 1.53 (1.44, 1.62) | 0.69 | 1.59 (1.47, 1.70) | 0.77 |
| Native T1 (ms) | 1,235 (1,222, 1,247) | 1,208 (1,197, 1,219) | 0.045 | 1,239 (1,222, 1,257) | 0.95 |
| ECV fraction (%) | 28.1 (27.0, 29.1) | 28.7 (27.7, 29.7) | 0.13 | 29.5 (28.0, 31.0) | 0.044 |
| Presence of LGE^δ^ (%) | 57.1 (38.2, 76.1) | 52.6 (36.3, 69.0) | 0.72 | 47.1 (22.2, 71.9) | 0.52 |
| ***LA Geometry ad Function*** |  |  |  |  |  |
| Indexed LA area (cm^2^/m^2^) | 11.2 (10.6, 11.9) | 12.4 (11.8, 13.0) | 0.016 | 11.5 (10.3, 12.7) | 0.67 |
| Minimum LAVi (mL/m^2^) | 33.4 (30.7, 36.0) | 36.9 (32.3, 41.4) | 0.16 | 38.2 (35.6, 40.8) | 0.029 |
| Maximum LAVi (mL/m^2^) | 13.8 (12.0, 15.6) | 14.8 (12.3, 17.3) | 0.56 | 16.0 (14.5, 17.5) | 0.056 |
| LAEF (%) | 59.1 (56.0, 62.2) | 58.2 (55.6, 60.8) | 0.63 | 60.3 (56.9, 63.6) | 0.61 |
| ***RV Geometry and Function*** |  |  |  |  |  |
| RVEDVi (mL/m^2^) | 77.0 (71.4, 82.5) | 83.0 (78.9, 87.0) | 0.099 | 87.0 (78.1, 95.8) | 0.029 |
| RVESVi (mL/m^2^) | 33.3 (29.8, 36.80 | 35.4 (33.0, 37.7) | 0.34 | 37.7 (32.5, 42.9) | 0.11 |
| RVEF (%) | 57.4 (55.0, 59.8) | 57.3 (55.4, 59.3) | 0.98 | 57.0 (54.3, 59.7) | 0.84 |
| TAPSE (cm) | 1.4 (1.3, 1.6) | 1.8 (1.6. 2.0) | 0.005 | 1.5 (1.2, 1.8) | 0.89 |

^α^ P value for differences in mean (or proportion^δ^) between HIV uninfected versus PHIV with early ART initiation.

^β^ P value for differences in mean (or proportion^δ^) between HIV uninfected versus PHIV with delayed ART initiation.

LV = left ventricle; LVM = LV mass; LVEDV = LV end diastolic volume; LVESV = LV end systolic volume; LVEF = LV ejection fraction; RWT = relative wall thickness; GCS = peak global circumferential strain; GLS = peak global longitudinal strain; GRS = peak global radial strain; DSR = peak diastolic strain rate; SIR = signal intensity ratio; ECV = extracellular volume; LGE = late gadolinium enhancement; LA = left atrium; LAV = LA volume; LAEF = LA ejection fraction; RV = right ventricle; RVEDV = RV end diastolic volume; RVESV = RV end systolic volume; RVEF = RV ejection fraction.

Postscript (i) = indexed to body surface area. rea.
